# Assessing Open Science practices in physical activity behaviour change intervention evaluations

**DOI:** 10.1101/2021.12.01.21267126

**Authors:** Emma Norris, Isra Sulevani, Ailbhe N. Finnerty, Oscar Castro

## Abstract

**Objectives:** Concerns on the lack of reproducibility and transparency in science have led to a range of research practice reforms, broadly referred to as ‘Open Science’. The extent that physical activity interventions are embedding Open Science practices is currently unknown. In this study, we randomly sampled 100 reports of recent physical activity behaviour change interventions to estimate the prevalence of Open Science practices.

**Methods:** One hundred reports of randomised controlled trial physical activity behaviour change interventions published between 2018-2021 were identified. Open Science practices were coded in identified reports, including: study pre-registration, protocol sharing, data-, materials- and analysis scripts-sharing, replication of a previous study, open access publication, funding sources and conflict of interest statements. Coding was performed by two independent researchers, with inter-rater reliability calculated using Krippendorff’s alpha.

**Results:** 78% of the 100 reports provided details of study pre-registration and 41% provided evidence of a published protocol. 4% provided accessible open data, 8% provided open materials and 1% provided open analysis scripts. 73% of reports were published as open access and no studies were described as replication attempts. 93% of reports declared their sources of funding and 88% provided conflicts of interest statements. A Krippendorff’s alpha of 0.73 was obtained across all coding.

**Conclusion:** Open data, materials, analysis and replication attempts are currently rare in physical activity behaviour change intervention reports, whereas funding source and conflict of interest declarations are common. Future physical activity research should increase the reproducibility of their methods and results by incorporating more Open Science practices.

## Introduction

Across scientific research, there is an increased awareness of highly prevalent problematic research practices, often referred to as Questionable Research Practices (QRPs) (Munafò et al., 2017). Such practices include *p-*hacking: trying numerous analysis approaches to find the most novel and significant results (Head et al., 2015; Simmons et al., 2011), cherry-picking: selective outcome reporting from large datasets (Mayo-Wilson et al., 2017) and Hypothesising After the Results are Known (or ‘HARKing’) (Murphy & Aguinis, 2019). Alongside this, a well-documented ‘replication crisis’ across science has shown that study findings can rarely be repeated in different contexts or even within the same contexts at a future timepoint (Ioannidis, 2005; Open Science Collaboration, 2015).

A global movement towards Open Science practices in research has arisen to address these QRPs. Open Science is an umbrella term representing a range of research behaviours intending to reduce QRPs, increasing the transparency and reproducibility of research (Munafò et al., 2018; Nosek et al., 2012). Open Science research practices can be applied across the whole research process: from conception to publication (FOSTER Open Science, 2021; Kathawalla et al., 2021). At research conception, pre-registrations (Field et al., 2020; Yamada, 2018) provide time-stamped evidence of study hypotheses, methods and analysis plans prior to data collection and make such information publicly available through online repositories such as [Open Science Framework](https://osf.io/) (Sullivan et al., 2019). Registered Reports provide in-principle acceptance to journals based on study proposals at conception stage, rather than based on completed studies and their reported findings (Chambers, 2019). Open data, open materials (including questionnaires and intervention materials used) and open analysis codes and scripts help make the processes and outputs of research more transparent, accessible and sharable (Nosek et al., 2015). At publication, Open Access publishing makes reporting of research available to anyone at no cost (Piwowar et al., 2018).

Various initiatives exist to facilitate and reward Open Science practices among researchers (Norris & O’Connor, 2019). For example, i) increased provision of free online training (Kizilcec et al., 2020); ii) journal badges recognising pre-registration, open data and open materials (Kidwell et al., 2016); iii) journal clubs to discuss and share Open Science practices (Orben, 2019); iv) discipline-specific organisations such as the Society for Improvement of Psychological Science (Steltenpohl et al., 2021) and v) national and international organisations, such as UK Reproducibility Network (Munafò et al., 2020) or German Reproducibility Network (Rahal et al., 2021): all aiming to increase the prevalence of Open Science practices across disciplines.

Meta-research has assessed Open Science practices in domains related to physical activity, behaviour change and life sciences. A recent study exploring 250 psychology studies of varying study designs published between 2014 and 2017 found that while open access publication was relatively common (65%), sharing of open materials (14%), data (2%) and analysis scripts (1%), as well as pre-specification of research plans via pre-registration (3%) and study protocols (0%) were low (Hardwicke et al., 2020). In addition, transparency of reporting was inconsistent for funding statements (62%) and conflict of interest disclosure statements (39%) (Hardwicke et al., 2020). Meta-science studies have also assessed these Open Science practices within smoking cessation behaviour change research (Norris et al., 2021), social sciences (Hardwicke et al., 2020), biomedicine (Wallach et al., 2018) and biostatistics (Rowhani-Farid & Barnett, 2018).

Recent calls to increase transparency in exercise and physical activity research are evident (Caldwell et al., 2020; Halperin et al., 2018). A recent study assessed the prevalence of questionable research practices within sport and exercise medicine research, including one hundred and twenty-nine studies published in leading sports medicine journals in 2019 (Büttner et al., 2020). Their analysis found that 82.2% of all reported hypotheses, and 70.8% of the primary hypothesis, were supported by study results, which authors identify as implausibly high (Büttner et al., 2020).

However, to our knowledge, no study has evaluated the extent to which Open Science practices are used within physical activity research. Gaining a better understanding of these practices could inform future recommendations and policy development to promote open, transparent science within the physical activity field and to reduce the threat of QRPs. Therefore, the aim of this current study is to assess Open Science practices within physical activity behaviour change intervention reports.

## Methods

### Study design

This was a retrospective observational study with a cross-sectional design. Sampling units were individual physical activity behaviour change intervention reports. This study applied a methodology used to assess reproducibility and transparency in smoking cessation interventions (Norris et al., 2021), psychological sciences (Hardwicke et al., 2021) and social sciences (Hardwicke et al., 2020).

This study was preregistered on the Open Science Framework:

https://doi.org/10.17605/OSF.IO/2ZYU3. All deviations from this protocol are explicitly acknowledged in Appendix 1.

### Search strategy

All papers included in this study were reports of physical activity behaviour change interventions, evaluated via randomised controlled trials. These reports were identified for inclusion within the [Human Behaviour-Change Project](https://www.humanbehaviourchange.org/) (Michie et al., 2017, 2020), which is developing an Artificial Intelligence system to extract information from published intervention studies and make recommendations for real-world practice and future research: https://osf.io/efp4x/. Physical activity behaviour change intervention reports were identified in this project using Microsoft Academic, one of the biggest, most comprehensive bibliographic databases of scientific literature (Visser et al., 2021). The search strategy was performed on 20.01.2021 and included the terms “MVPA or moderate-to-vigorous physical activity or MPA or VPA or moderate physical activity or vigorous physical activity or strenuous physical activity or hard physical activity”, with studies additionally filtered using the Randomised Controlled Trial classifier (https://academic.microsoft.com/topic/168563851).

Inclusion criteria were reports describing randomised controlled trials of physical activity behaviour change interventions and published between 2018 and 2021. The rationale for the recency of these included papers is to best represent current Open Science practices, given the relatively recent nature of Open Science practices (Munafò et al., 2017). In addition, we focused on randomised controlled trials only due to their recognition as ‘gold-standard’ for studying intervention effectiveness (Michie et al., 2017). Exclusion criteria were trial protocols, abstract-only entries, qualitative research and economic or process evaluations. Titles and abstracts of identified papers within the Human Behaviour Change Project were screened by one researcher (EN) to double-check relevance against inclusion and exclusion criteria. Of the 181 reports remaining after applying these criteria, 100 reports were selected using the [Calculator Soup Random Number Generator](https://www.calculatorsoup.com/calculators/statistics/random-number-generator.php).

### Measures

Article characteristics recorded were: i) author name, ii) publication year and iii) country of the corresponding author. Open Science practices were assessed by recording presence of the following in included reports: i) *Pre-registration:* whether pre-registration was reported as carried out, where the pre-registration was hosted (e.g Open Science Framework, ClinicalTrials.gov), whether it could be accessed, what aspects of the study (hypotheses, methods and analysis plans) were pre-registered and whether the pre-registration was logged prospectively (prior to data collection commencing) or retrospectively (after data collection had commenced) (Loder et al., 2018); ii) *Protocol sharing:* whether a protocol was reported as published and what aspects of the study (hypotheses, methods and analysis plans) were included in the protocol; iii) *Data sharing*: whether data were reported as available, where it was available (e.g online repository such as Open Science Framework, upon request from authors, as a journal supplementary file), whether the data were downloadable and accessible, whether data files were clearly documented and the extent that data reported were sufficient to allow replication of study findings; iv) *Materials sharing:* whether study materials were reported as available, where they were available (e.g online repository such as Open Science Framework, upon request from authors, as a journal supplementary file) and whether the materials were downloadable and accessible; v) *Analysis script-sharing:* whether analysis scripts were reported as available, where they were available (e.g online repository such as Open Science Framework, upon request from authors, as a journal supplementary file) and whether the analysis scripts were downloadable and accessible; vi) *Replication of a previous study:* whether the study was described as being a replication attempt of a previous study; vii) *Open access publication:* whether the study was published as open access, assessed via the [Open Access button](https://openaccessbutton.org/) website; viii) *Funding sources:* whether funding sources were declared and if research was funded by public organisations (such as research councils or charities), pharmaceutical, activity-related or other companies; and ix) *Conflicts of interest:* whether conflicts of interest were declared and whether conflicts were with public organisations (such as research councils or charities), pharmaceutical, activity-related or other companies. All measured variables are shown in Table 1.

**Table 1.** Measured characteristics within identified physical activity behaviour change intervention reports.

| Variables | Coder questions | Response options |
| --- | --- | --- |
| **Article characteristics**<br>**Coder instructions:** Check the institutional affiliation of the corresponding author. If there are multiple corresponding authors, choose the first. If no corresponding author is identified, choose the first. If there are multiple affiliations for the selected author, choose the first. |  |  |
| Country | Which country is the corresponding author based in according to their affiliated? | [list countries]/ Unclear/ Other |
| **Pre-registration**<br>**Definitions:** “Pre-registration” refers to the specification of important aspects of the study (typically hypotheses, methods, and/or analysis plan) prior to commencement of the study.<br>**Coder instructions:** Check specific sections in the paper where these files might be located e.g., supplementary materials, appendices, author notes, methods, and results sections. Search for “registration”. |  |  |
| Pre-registration statement | Does the article state whether or not the study (or some aspect of the study) was pre-registered? | Yes – the statement says that there was a pre-registration /<br>Yes – the statement says that there was NO pre-registration /<br>No – there is no pre-registration statement / Other* |
| Pre-registration method | Where does the article indicate the pre-registration is located? | Open Science Framework /<br>AsPredicted /<br>ClinicalTrials.gov /<br>AEA Trial Registry /<br>EGAP Registry /<br>Registered Report / Other* |
| Pre-registration accessible | Can you access and open the pre-registration | Yes / No / Other* |
| Pre-registration content | What aspects of the study appear to be pre-registered? (select all that apply) | Hypotheses<br>Methods<br>Analysis Plan<br>Other* |
| **Protocol sharing**<br>**Definition:** “protocol” refers to a document containing details about the study design, methods, and analysis plan. It may or may not be pre-registered.<br>**Coder instructions:** Search the article for the phrase ‘protocol’ and assess whether a link is provided to a protocol document. |  |  |
| Protocol availability | Does the article link to an accessible protocol? | Yes / No / Other* |
| Protocol content | What aspects of the study appear to be included in the protocol? (select all that apply) | Hypotheses<br>Methods<br>Analysis Plan<br>Other* |
| **Data sharing**<br>**Definitions:** “data” refers to recorded information that supports the analyses reported in the article. A “data availability statement” can be as simple as a url link to a data file, or as complex as a written explanation as to why data cannot be shared. |  |  |

| **Coder instructions:** Check the article for a data availability statement/link. They are often located in the "supplementary material", "acknowledgements", "author notes", "methods", or "results" sections. Search the article for the text "data availab" (to cover "data availability" and "data available"). |  |  |
| --- | --- | --- |
| Data availability statement | Does the article state whether or not data are available? | Yes - the statement says that the data (or some of the data) are available /<br>Yes - the statement says that the data are NOT available /<br>No - there is no data availability statement / Other* |
| Data sharing method | How does the statement indicate the data are available? | Upon request from the authors / Personal or institution website /<br>An online, third-party repository (e.g., OSF, FigShare etc.) /<br>supplementary materials hosted by the journal / Other* |
| Data accessibility | Can you access, download, and open the data files? | Yes / No / Other* |
| Data documentation | Are the data files clearly documented? | Yes / No / Other* |
| Data content | Do the data files appear to contain all of the raw data necessary to reproduce the reported findings? | Yes / No / Unclear / Other* |
| **Materials sharing**<br>**Definitions:** "materials" refers to any study items that would be needed to repeat the study, such as stimuli, survey instruments, and computer code/software used for data collection, presentation stimuli or running experiments |  |  |
| Materials availability statement | Does the article state whether or not materials are available? | Yes - the statement says that the materials (or some of the materials) are available /<br>Yes - the statement says that the materials are NOT available /<br>No - there is no materials availability statement / Other* |
| Materials sharing method | According to the statement, how are the materials accessible? | Upon request from the authors / Personal or institution website /<br>An online, third-party repository (e.g., OSF, FigShare etc.) /<br>supplementary materials hosted by the journal / Other* |
| Materials accessibility | Can you access, download, and open the materials files? | Yes / No / Other* |

| **Analysis script sharing**<br>**Definition:** "Analysis scripts" refers to specification of data preparation and analysis steps in the form of highly detail step-by-step instructions for using point-and-click software, analysis code (e.g., R), or syntax (e.g., from SPSS).<br>**Coder instructions:** Check the article for an analysis script availability statement/link. They are often located in the "supplementary material", "acknowledgements", "author notes", "methods", or "results" sections. Search for the text "analysis script" and "analysis code". |  |  |
| --- | --- | --- |
| Analysis script availability statement | Does the article state whether or not analysis scripts are available? | Yes - the statement says that the analysis scripts (or some of the analysis scripts) are available /<br>Yes - the statement says that the analysis scripts are NOT available /<br>No - there is no analysis script availability statement |
| Analysis script sharing method | According to the statement, how are the analysis scripts accessible? | Upon request from the authors / Personal or institution website /<br>An online, third-party repository (e.g., OSF, FigShare etc.) /<br>supplementary materials hosted by the journal /<br>Other* |
| Analysis script accessibility | Can you access, download, and open the analysis script files? | Yes / No / Other* |
| **Funding**<br>**Coder instructions:** Funding is usually reported in a specific section e.g., "Author information", or "Funding statement". Search the article for the phrase "funding". If you are unsure whether an organisation is an activity-related company, pharmaceutical company, other private company or public organisation, Google the organisation name and code accordingly. If it is unclear to you whether the funding is private or public, choose the 'other' option and enter 'unclear'. |  |  |
| Funding statement | Does the article include a statement indicating whether there were funding sources? | Yes – the statement says that there was funding from an activity-related company (e.g Nike, Fitbit) /<br>Yes – funding from a pharmaceutical company (e.g Pfizer, GSK)/<br>Yes – funding from another private company (e.g Google, Coca Cola) /<br>Yes – funding from a public organisation (e.g National Institute of Health Research)/<br>Yes - the statement says that there was no funding was provided /<br>No – there is no funding statement /<br>Unclear/Other* |

| Conflict of interest |  |  |
| --- | --- | --- |
| **Coder instructions:** Conflicts of interest are usually reported in a specific section e.g. “Author information” or “Conflict of interest statement”. Search the article for the phrases “conflict of interest” and/or “competing interest”. If you are unsure whether an organisation is an activity-related company, pharmaceutical company, other private company or public organisation, Google the organisation name and code accordingly. If it is unclear to you whether the conflict of interest is private or public, choose the 'other' option and enter 'unclear'. |  |  |
| Conflict of Interest statement | Does the article include a statement indicating whether there were any conflicts of interest? | Yes – the statement says that there was a conflict of interest from an activity-related company (e.g Nike, Fitbit) /<br>Yes – conflict of interest from a pharmaceutical company (e.g Pfizer, GSK)/ /<br>Yes – conflict of interest from another private company (e.g Google, Coca Cola) /<br>Yes – conflict of interest from a public organisation (e.g National Institute of Health Research)/<br>Yes - the statement says that there is no conflict of interest /<br>No – there is no conflict of interest statement /<br>Other* |
| **Replication**<br>**Definitions:** “replication” refers to repetition of a previous study’s methods in order to ascertain whether similar findings can be obtained.<br>**Coder instructions:** Search the abstract and introduction for the phrase “replicat” (to cover ‘replication’, ‘replicates’ etc). Confirm the authors are using the phrase with the definition provided above. |  |  |
| Replication statement | Does the article claim to report a replication study? | The article claims to report a replication study (or studies) /<br>There is no clear statement that the article reports a replication study (or studies) /<br>Other* |
| **Open access**<br>**Coder instructions:** To establish the open access status of the article: Go to https://openaccessbutton.org/ and enter the article’s doi (e.g., “10.1371/journal.pcbi.1004574”) if available (if not, enter the article title). If a link is provided, check that you can access the article at the link. If the article is accessible answer “Yes”. If the article is not accessible at the provided link, or no link is provided, answer “No”. |  |  |
| Open access status | Is the article open access? | Yes – found via open access button / Yes – found via other means / No – could not access article other than through paywall / Other* |

\*If a response marked with an asterisk is selected, the coder is asked to provide more detail in a free text response box.

### Procedure

Coding of identified intervention reports took place between July and September 2021, with all data extracted onto a Google Form (https://osf.io/3wcu6/). All reports were independently coded by two researchers (IS coded all 100 papers, EN & OC coded 50 each). Any discrepancies were resolved through discussion, with input from a third researcher not involved in the initial coding if required (EN or OC).

## Analysis

Raw numbers and percentages were identified for each variable. Inter-rater reliability of the independent coding by the two researchers was calculated using Krippendorff’s alpha (Hayes & Krippendorff, 2007) using R package ‘irr’ version 0.84.1 https://osf.io/e7f84/.

## Results

### Sample characteristics

Twenty-two out of the 100 physical activity behaviour change intervention reports were published in 2018, thirty-three in 2019, thirty-seven in 2020 and eight in 2021. The 100 reports evaluated studies conducted in twenty-four different countries, taking place most commonly in the United States of America (k=24), Australia (k=19), Canada (k=10) and the United Kingdom (k=7). A full summary of countries in included reports is presented [here](https://osf.io/cwnrx/).

### Open Science practices in physical activity behaviour change intervention reports

Final reconciled coding of Open Science practices for all 100 included physical activity behaviour change intervention reports can be found [here](https://osf.io/cwnrx/).

### Article availability (Open Access)

Seventy-three out of 100 physical activity behaviour change intervention reports were available via open access, with twenty-seven of them only accessible behind a paywall (Figure 1A).

**Figure 1A - 1I.**
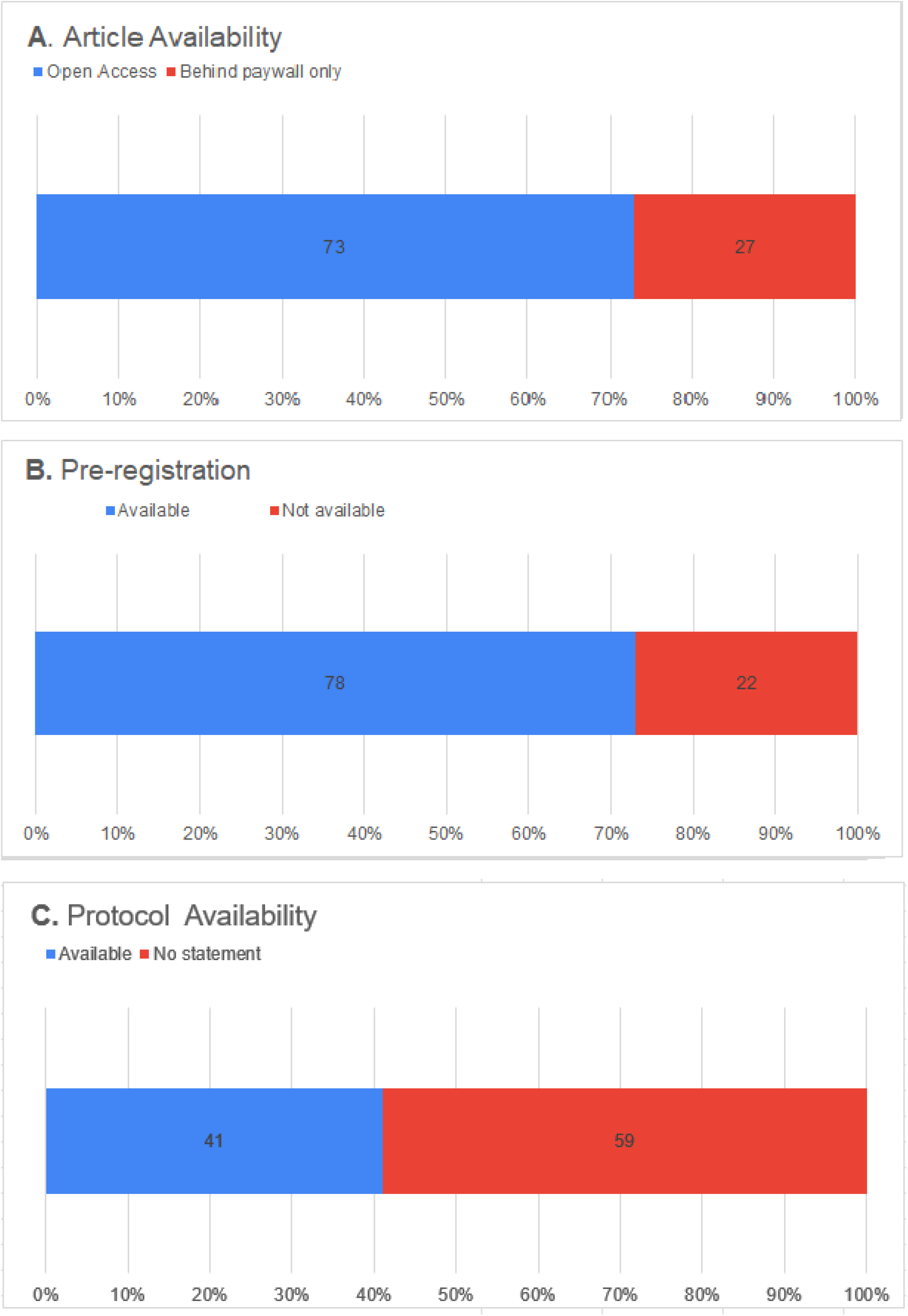
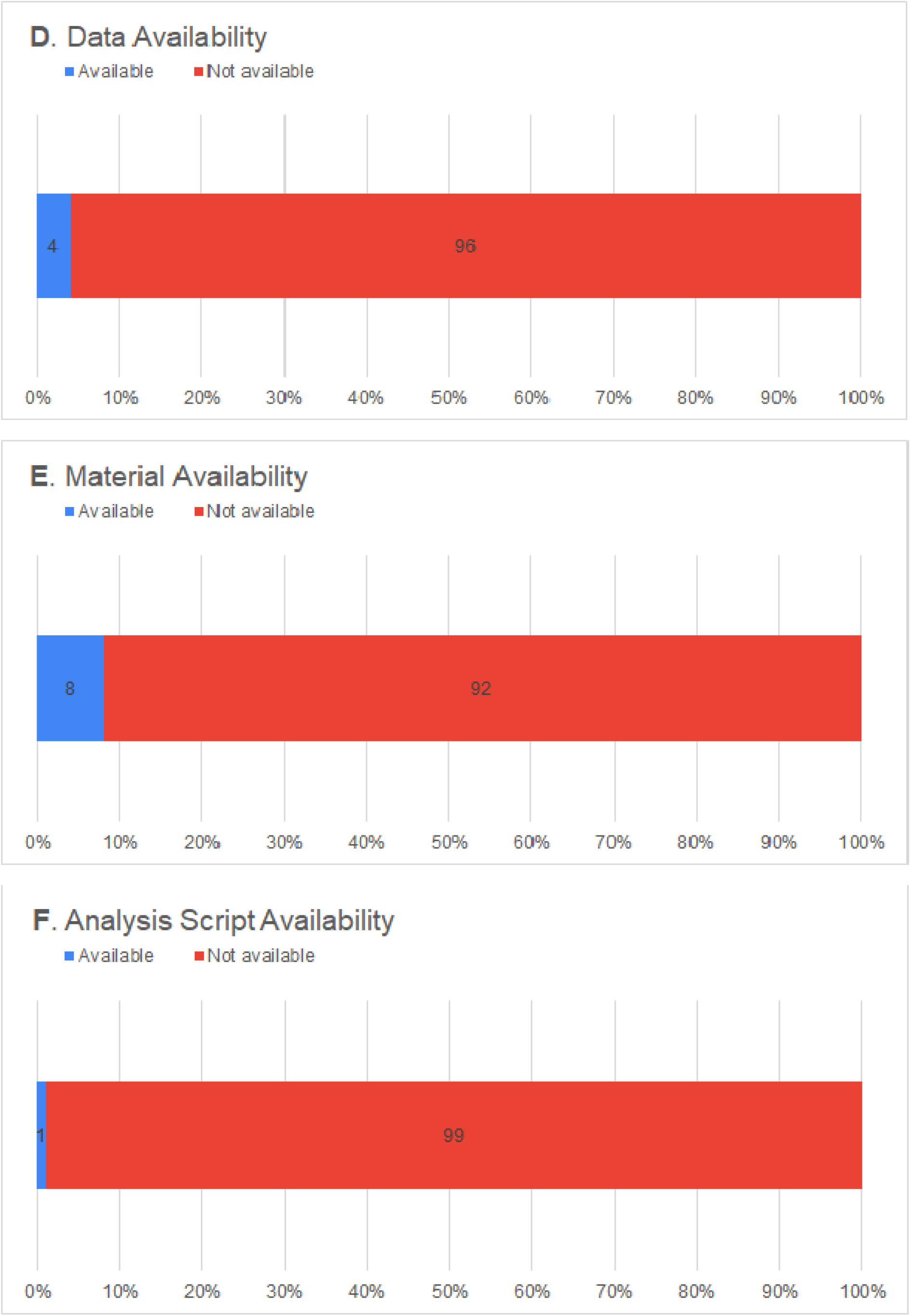
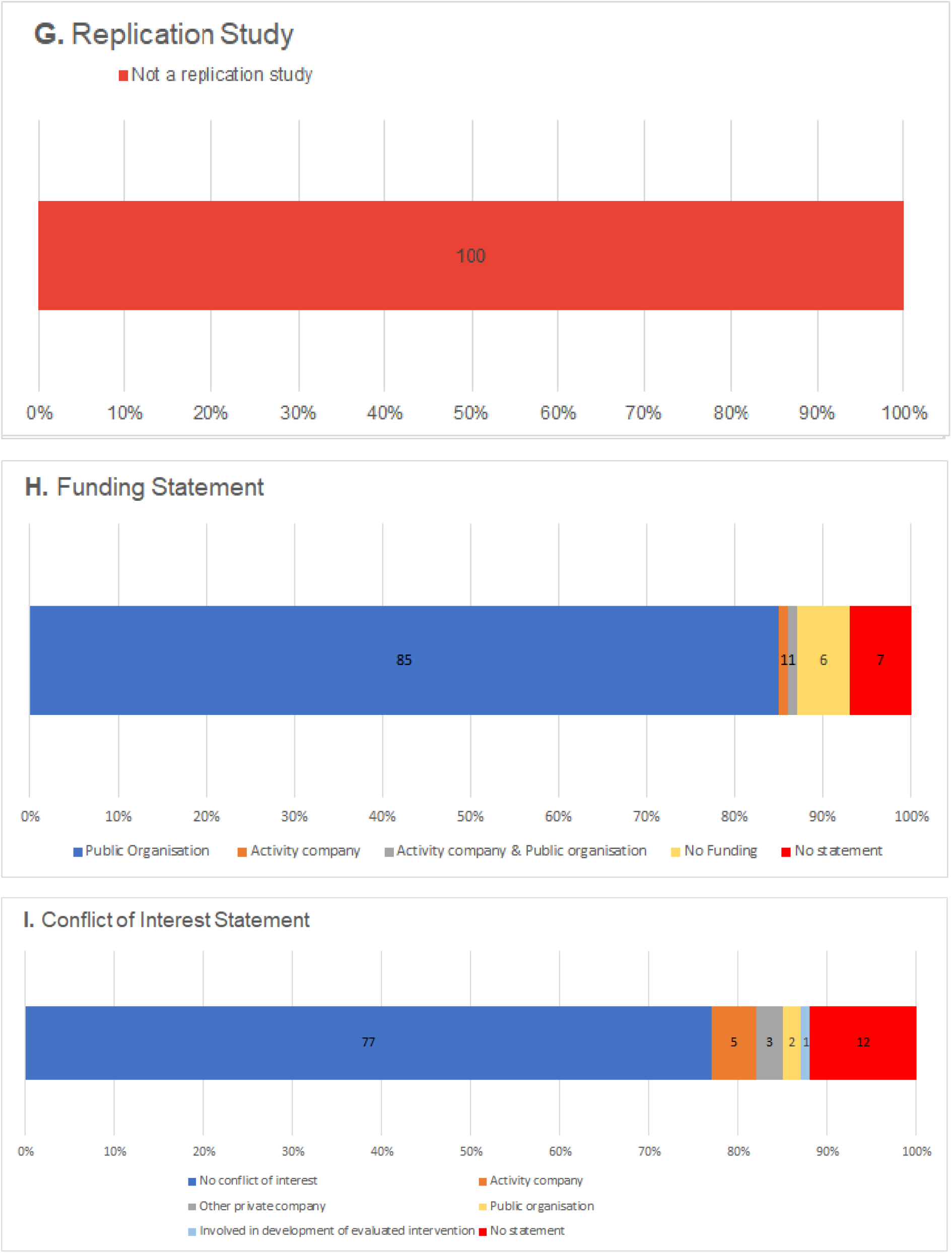
Open Science practices in physical activity behaviour change intervention reports.

### Pre-registration

Seventy-eight out of 100 physical activity behaviour change intervention reports included a statement indicating existence of a study pre-registration. Of those, seventy-seven could be accessed. Forty-three of all accessible pre-registrations were recorded prospectively (i.e before data collection commenced) and thirty-four were recorded respectively (i.e after data collection can commenced). Seventy-seven of all accessible pre-registrations declared specifications relating to study methods, twenty-four declared hypotheses and five declared analysis plans. Thirty-seven of all accessible pre-registrations were hosted on ClinicalTrials.gov (48.1%), eighteen on the Australian and New Zealand Clinical Trials Registry ([ANZCTR](https://www.anzctr.org.au/): 23.4%), fourteen on the International Standard Randomized Clinical Trial Number registry ([ISRCTN](https://www.isrctn.com/): 18.2%), three on Netherlands Trial Register ([NTR](https://www.trialregister.nl/api): 3.9%) and one on Deutsches Register Klinischer Studien ([DRKS](https://www.drks.de/drks_web/)), Iranian Registry of Clinical Trials ([IRCT](https://www.irct.ir)), Registro Brasileiro de Ensaios Clinicos ([REBEC](https://ensaiosclinicos.gov.br)), UMIN Clinical Trials Registry ([UMIN-CTR](https://www.umin.ac.jp/ctr/)) and Chinese Clinical Trial Registry ([ChCTR](http://www.chictr.org.cn/enindex.aspx)) (1.3% each). One included study was noted as a Registered Report (Ek et al., 2020), logged with an International Registered Report Identifier ([IRRID](https://support.jmir.org/hc/en-us/articles/360003797672-What-is-an-International-Registered-Report-Identifier-IRRID): 1.3%) (Figure 1B).

### Protocol sharing

Forty-one out of 100 physical activity behaviour change intervention reports included a statement about protocol availability. All forty-one (100%) of these protocols specified study methods, forty (97.6%) specified analysis plans and fourteen (34.1%) specified hypotheses (Figure 1C).

### Data sharing

Thirty-two out of 100 physical activity behaviour change intervention reports included a data availability statement. Of those, twenty-two stated data was only available upon request from the authors, five stated that data was available within the reports’ supplementary files, one stated that data was available via a personal or institutional website, and four stated that data was not available. Only four out of these thirty-two reports included a data availability statement that data files that were actually accessible to download, with two of these providing clear documentation for the data files and two providing sufficient detail needed to reproduce findings (Figure 1D).

### Material sharing

Seventeen out of 100 physical activity behaviour change intervention reports included a materials availability statement. Of those, ten reports stated that materials were available within the reports’ supplementary files and seven stated that materials were only available upon request from the authors. Eight out of the ten studies which stated that materials were provided as supplementary files actually provided accessible and downloadable materials, such as full or sample intervention activities (Figure 1E).

### Analysis script sharing

One out of 100 physical activity behaviour change intervention reports included an analysis script availability statement (Tudor-Locke et al., 2020), with this provided as a supplementary file (Figure 1F).

### Replication study

None of the 100 physical activity behaviour change intervention reports were described as replication studies (Figure 1G).

## Data Availability

All data from this study are available on Open Science Framework.

https://osf.io/t5gw4/

## Inter-rater reliability assessment

Inter-rater reliability of all coding across the 100 reports was assessed as good, *a*=0.73.

## Discussion

This study aimed to assess Open Science practices within physical activity behaviour change intervention reports. It was found that Open Science practices varied amongst the assessed 100 physical activity behaviour change intervention reports. Most reports were open access and pre-registered, with reported funding sources and conflicts of interest. However, research materials, data and analysis scripts were not frequently provided and no replication studies were identified.

Pre-registration of studies was found to be slightly more common for physical activity intervention RCTs (78%), than found in smoking cessation intervention RCTs (73%: Norris et al., 2021) and much more common than in wider psychological research of varying study designs (3%) (Hardwicke et al., 2021). In our study, similar amounts of studies were preregistered prospectively (55.7%: prior to data collection commencing) or retrospectively (44.2%: after data collection had commenced; Loder et al., 2018), although this distinction between preregistrations has not been assessed in comparable research. The common prevalence of retrospective pre-registration via clinical trials is arguably not true pre-registration, nor transparent from the study’s outset (Field et al., 2020; O’Connor, 2021; Yamada, 2018). One included study was noted as a Registered Report (Ek et al., 2020), where in-principle acceptance to journals is given based on study proposals at conception stage, rather than based on completed studies and their reported findings (Chambers, 2019). No Registered Reports were identified in smoking cessation (Norris et al., 2021) and psychology (Hardwicke et al., 2021), perhaps reflecting a slow increase in Registered Report numbers over time (Chambers, 2019; Scheel et al., 2021). Protocols were available as separate papers or linked publications in 41% of included physical activity studies, which is higher than in smoking cessation studies (29%; Norris et al., 2021) and wider psychology research (0%; Hardwicke et al., 2021). The increased prevalence of protocols within physical activity and smoking cessation likely reflects greater availability of health-related protocol publications (Basu et al., 2017), via specific journals such as [JMIR Research Protocols](https://www.researchprotocols.org/) and via protocols as specific types of publications within wider journals such as [BMC Public Health](https://bmcpublichealth.biomedcentral.com/submission-guidelines/preparing-your-manuscript/study-protocol) and [Trials](https://trialsjournal.biomedcentral.com/about?gclid=Cj0KCQjw5oiMBhDtARIsAJi0qk2VWOadbO06TIRd8XsTeyVk0UJVKTPrvccDFpbyvzVMNcW3sa6EhyQaAkKcEALw_wcB). High prevalence of protocols in this study is also indicative of randomised controlled trials being both a common study design in health and intervention research (Deaton & Cartwright, 2018) and a study design typically accompanied by research protocols (Tetzlaff et al., 2012).

Open access reports were at similarly moderate levels in physical activity (73%) than in smoking cessation (71%; Norris et al., 2021) and psychology (65%; (Hardwicke et al., 2021), but greater than the 45% observed in the social sciences (Hardwicke et al., 2020), the 45% across scientific literature published in 2015 (Piwowar et al., 2018) and the 25% in biomedicine (Wallach et al., 2018). This high rate of open access publishing in physical activity interventions may reflect increasing requirements by health funding bodies for open access publications (Severin et al., 2020), as well as increasing usage of preprint servers such as [MedRxiv](https://www.medrxiv.org/) for medical sciences and [PsyArXiv](https://psyarxiv.com/) for the psychological sciences (Flanagin et al., 2020; Peiperl & Editors, 2018).

Open materials were less commonly available in physical activity reports (8%) than in smoking cessation reports (13%; (Norris et al., 2021), psychology (14%; Hardwicke et al., 2021) and biomedicine (33%; Wallach et al., 2018). Open data was also less common across physical activity reports (4%) than in smoking cessation reports (7%; Norris et al., 2021), but greater than the 2% of wider psychological research (Hardwicke et al., 2021). Provision of raw data as supplementary files to published intervention reports or via trusted third-party repositories such as the [Open Science Framework](https://osf.io/) (Klein et al., 2018) is important to facilitate evidence synthesis. Open analysis scripts were found to be as infrequently provided in physical activity studies than in smoking interventions and wider psychological research (all 1%) (Hardwicke et al., 2021; Norris et al., 2021). No replication attempts were identified in this sample of physical activity intervention reports, same as within smoking cessation reports (Norris et al., 2021) but less than in the social sciences (1%; Hardwicke et al., 2020) and in wider psychology studies (5%; Hardwicke et al., 2021).

Declaration of funding sources were declared in physical activity reports (93%) similarly to smoking cessation reports (95%; Norris et al., 2021), more so than wider psychology (62%; Hardwicke et al., 2021), social sciences (31%; Hardwicke et al., 2020) and biomedical science reports (69%; Wallach et al., 2018). Similarly, a conflict of interest statement was provided as commonly in physical activity reports than in smoking cessation reports (88% in both; Norris et al., 2021) and higher than in wider psychology (39%; Hardwicke et al., 2021), social sciences (39%; Hardwicke et al., 2020) and biomedical sciences reports (65%; Wallach et al., 2018). Eight percent of studies reported conflicts from private companies including activity, pharmaceutical and other companies, less than the 20% of studies reporting company funding in smoking cessation interventions (Norris et al., 2021).

### Future steps to increase Open Science in physical activity interventions

This research has demonstrated a need to address the low levels of Open Science engagement in physical activity research, particularly in the areas of open materials, data, analysis scripts and replication attempts. As with any complex behaviour change, this transformation requires systems change across bodies involved in the development, running and publication of physical activity research: researchers, research institutions, funding organisations, journals and beyond (McVay & Conroy, 2021; Munafò et al., 2017). In order to develop effective behaviour change interventions, it is important to use a systematic and comprehensive approach to intervention development, underpinned by a model of behaviour and theoretically predicted mechanisms of action (Michie & Johnston, 2012; Willmott & Rundle-Thiele, 2021). The Capability, Opportunity, Motivation, Behaviour (COM-B) model (Michie et al., 2011) posits that changing behaviour involves changing one or more of the following: capability (psychological and physical capacity to engage in the behaviour), opportunity (external factors that make the execution of a particular behaviour possible or prompt it) and motivation (internal processes that energise and direct behaviour). We argue that understanding the capability, opportunity and motivation associated with Open Science practices (Norris & O’Connor, 2019; O’Connor, 2021) and developing interventions to address these determinants of behaviour change, is key to increase engagement with Open Science.

For example, low perceived capability towards Open Science practices in physical activity researchers can be addressed by providing researchers with training tailored to the context of activity intervention research (e.g., online training on how to make anonymised activity monitor data openly available, how to use preprint servers most relevant to activity research or how to make their activity analysis reproducible). Opportunity to engage in Open Science practices can be facilitated within institutions, encouraging discussions around Open Science in the context of physical activity research (Büttner et al., 2020) and in science more broadly (Munafò et al., 2020; Orben, 2019), as well as developing a research culture valuing and promoting the benefits of open science practices (Munafò et al., 2020; Nosek et al., 2015). Motivation for Open Science can be addressed by providing incentives, such as awarding funding to research-embedding open practices (Smaldino et al., 2019).

Similarly, Open Science badges recognising open data, materials and pre-registration have been adopted by journals as a simple, low-cost scheme to reward these research behaviours (Kidwell et al., 2016; Rowhani-Farid & Barnett, 2018). However, uptake of Open Science badges in physical activity journals is currently low and is rife for increased uptake in the field.

### Strengths and limitations

A strength of this study is the implementation of a comprehensive and previously used approach to identify open science practices. Moreover, two researchers independently carried out the coding of open science practices, reducing the risk of human error and maximising reliability (Gwet, 2014). A limitation is that results are based on a relatively small sample of physical activity behaviour change reports, meaning findings may not be applicable to all physical activity research. In addition, the assessment of open science practices was entirely dependent on what was described within evaluation reports. Direct requests to authors or additional wider searching of third-party registries such as Open Science Framework may have identified additional information.

## Conclusions

Open Science practices in physical activity behaviour change intervention reports were varied. Open access publication and pre-registration of research plans were common, although pre-registration was often done retrospectively i.e after data collection has started, hence not in the most transparent manner. Provision of open data, materials and analysis was rare and replication attempts were non-existent. Funding sources and conflicts of interest were usually declared. Urgent initiatives are needed to increase the uptake of all Open Science practices in physical activity, with a particular focus on open materials, data, analysis scripts and replication attempts.

## Statements

### Contributorship statement

Emma Norris: Conceptualisation, data curation, formal analysis, investigation, methodology, project administration, resources, software, supervision, validation, visualisation, writing – original draft, writing – review & editing; Isra Sulevani: Data curation, formal analysis, writing – review & editing; Ailbhe N. Finnerty: Formal analysis, software, writing – original draft, writing – review & editing; Oscar Castro: Conceptualisation, data curation, formal analysis, investigation, methodology, writing – original draft, writing – review & editing.

### Data sharing statement

All data from this study are available here: https://osf.io/t5gw4/

### Ethical approval

Not required as this was meta-research of existing research.

## Appendix 1. Updates to pre-registration

During the course of this study and peer review, we made minor adjustments to the pre-registered protocol:

1. We added an additional item: ‘Was the declared pre-registration actually pre-registered ahead of data collection?’ (Question 8). This specified whether a declared study pre-registration was actually pre-registered ahead of data collection, or whether it was actually retrospectively registered after data collection had commenced. Responses for this new item were ‘Yes – this pre-registration was registered ahead of data collection’ or ‘No – this pre-registration was retrospectively registered after data collection had commenced’
2. The presence of ‘supplementary material’ was not sufficient for Material availability alone, as supplementary materials feature a variety of documents from protocols, materials, data etc.
3. Where a data and/or materials availability sub-heading was present in a paper, but no discussion of data and/or materials availability was actually given there; we coded the paper as ‘No - there is no data availability statement’/’No - there is no materials availability statement’.
4. We added an option ‘Yes - Funded by a non-profit’ under Funding sources.
5. We added an option ‘Yes - Funded by a non-profit’ under Conflict of Interest.
6. We added an option ‘Yes - Statement says researchers involved in both development and evaluation of intervention’ under Conflict of Interest.

